# On nonlinear incidence rate of Covid-19

**DOI:** 10.1101/2020.10.19.20215665

**Authors:** Swarna Kamal Paul, Saikat Jana, Parama Bhaumik

**Affiliations:** Tata Consultancy Services, Gitanjali Park, Newtow, Kolkata, India; Parama Bhaumik is with the Information Technology Department, Jadav- pur university, Saltlake, Kolkata, India

**Keywords:** Artificial Intelligence, Discrete Mathematics, Neural nets, modeling and prediction

## Abstract

Classical Susceptible-Infected-Removed model with constant transmission rate and removal rate may not capture real world dynamics of epidemic due to complex influence of multiple external factors on the spread. On top of that transmission rate may vary widely in a large region due to non-stationarity of spatial features which poses difficulty in creating a global model. We modified discrete global Susceptible-Infected-Removed model by using time varying transmission rate, recovery rate and multiple spatially local models. No specific functional form of transmission rate has been assumed. We have derived the criteria for disease-free equilibrium within a specific time period. A single Convolutional LSTM model is created and trained to map multiple spatiotemporal features to transmission rate. The model achieved 8.39% mean absolute percent error in terms of cumulative infection cases in each locality in a 10-day prediction period. Local interpretations of the model using perturbation method reveals local influence of different features on transmission rate which in turn is used to generate a set of generalized global interpretations. A what-if scenario with modified recovery rate illustrates rapid dampening of the spread when forecasted with the trained model. A comparative study with current normal scenario reveals key necessary steps to reach baseline.

## 1 Introduction

Dynamical systems equations based on compartmental modelling of epidemiology have been widely used to predict the spread of an epidemic. Susceptible-Infected-Removed or SIR model is one such simplified set of differential equations to model the spread. However, accurately determining parameter values like the transmission rate for a specific disease is a challenge. The dynamics of a disease may vary across space and time. Many external factors may influence the transmission rate. Considering the transmission rate constant for a disease, grossly oversimplifies the model, thus compromising accuracy. Secondly, knowing the factors influencing the transmission rate and the dynamics of the influence can provide a vivid understanding of the disease progression.

There are several different types of nonlinear incidence rate suggested in the literature [3,4, 5, 6,7,8,9]. However, most of them adopt some type of simple predefined function with few parameters to model the incidence rate. Simple functions have low representational capability. Thus, they may not capture the detail dynamical variations of the incidence rate caused by multiple factors. We propose a Convolutional LSTM based spatiotemporal model to map the transmission rate of Covid-19 with respect to multiple input features and thereby map the derived incidence rate from transmission rate. The model can forecast incidence rate with high spatiotemporal resolution provided availability of clean historical data in that resolution. Exploratory analysis reveals probable influence of external features on transmission rate and eventually helped in feature selection. A spatiotemporal local interpretation method of a black box model is proposed which in turn is used to explain the trained model. The explanations reveal local influence of different external features on the transmission rate. A generalized global explanation is also generated to find common influence of factors across multiple locations and over a period.

We experimented with available data of Covid-19 across multiple regions of USA and the model achieved 7.95% and 0.19% mean absolute percent error in terms of new infection cases in each locality and cumulative total infection cases across the country in a 10-day prediction period respectively. The generated explanations revealed high influence of population density, somewhat medium influence of gender ratio and median population age on the transmission rate, globally. There are minor influences of temperature and temperature deviation but barely any observable influence of humidity. However, local influences of features vary widely across multiple small regions. A criterion for disease-free equilibrium within a specific time period has been derived for discrete SIR model with variable transmission and recovery rate. A long-term forecast using the trained model and modified recovery rate to satisfy disease-free equilibrium criteria reveals rapid damping of active infection cases to reach the baseline. However frequent spikes due to resurgence are seen in this scenario. A comparative study is made with forecasted dynamics using current normal recovery rate to reveal necessary actions for rapid containment of the disease.

The paper is organized as following. We conducted a brief literature survey in section 2. Section 3 briefly explains the discrete SIR model with variable transmission rate. Section 4 discusses about spatiotemporal modelling of transmission rate. Section 5 discusses on spatiotemporal influence of external features on transmission rate. We conduct long term forecasting of disease progression with a current normal scenario and a “what-if” scenario in section 6. Section 7 concludes the paper.

## 2 Related work

Kermack and Mckendrick [1] modelled communicable diseases using differential equations. Hethcote introduced the SIR model [2] where population is compartmentalized into susceptible, infected and removed groups. A set of differential equations modeled the dynamics of population in different compartments. In traditional SIR model incidence rate or the number of new infections per unit time varies bilinearly with the number of infections and number of susceptible in a population considering the transmission rate as constant. However, assumptions like homogenous mixing, non-dependence on external factors, no psychological effects on population etc. may not be realistic in many cases. Thus, several authors [3,4, 5, 6,7,8,9] introduced different types of non-linear incidence rates mostly addressing the saturation and psychological effect. Saturation effect states that the incidence rate might slow down and saturate as number of infected individuals increases due to low availability of susceptible individuals. Psychological effect on the population results in increased cautiousness among susceptible individuals as the epidemic spreads thus, slowing down the transmission rate. Most of the incidence rates stated above satisfy weakly non-linear property and are too simple to capture any arbitrary effects of the environment. SIR model with time varying transmission recovery rate have been studied in [11] and thresholds theorems are derived. Liu et. al. [10] introduced a time varying switched transmission rate to model nonlinear incidence.

Hu et. al. developed a modified stacked autoencoder model of the epidemic spread in China and they claimed to achieve high level of forecasting accuracy [26]. On observing a universality in the epidemic spread in each country, Fanelli and Piazza [27] applied mean-field kinetics of Susceptible-Infected-Recovered/Dead epidemic model to forecast the spread and provided an estimation of peak infections in Italy. Zhan et. al. [14] integrated the intercity migration data in China with Susceptible-Exposed-Infected-Removed model to forecast an estimation of epidemic spread in China. Hong et. al. [12] considered variable transmission rate of Covid19 and came up with variable Rnaught factor of Covid-19. Xi et. al. [28] used deep residual networks to model spatiotemporal characteristics of the spread of influenza and experimented with real dataset of Shenzhen city in China. Paul et. al. [35] used ensemble of ConvLSTM networks to forecast Covid-19 total infection cases.

## 3 Discrete sir model with variable transmission rate

In SIR model the total population in a region is compartmentalized into 3 classes, namely Susceptible (S), Infected (I) and Removed (R). Initially the whole population is in susceptible class. An individual can move from susceptible to infected class on contracting the disease. An infected individual can move to removed class by either getting recovered and immune to the disease or deceased. The dynamics of the disease spread can be modelled by the following set of differential equations.

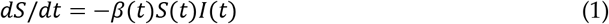

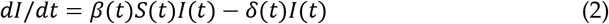

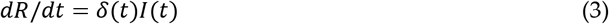

Where *β*(*t*) is disease transmission rate or contact rate and *δ*(*t*) is removal rate which is sum of recovery rate and mortality rate. It is assumed the population size (*N*) remains constant during the course of epidemic. *S*(*t*), *I*(*t*) and *R*(*t*) are scaled as fraction of total population. Thus, the following equation holds true.

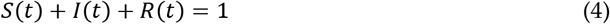

From [11] we get the following ∀*t* > *t*_0_, where *r* = *R*(*t*_0_), *R*(*t*) ≥ *r* ∀*t* > *t*_0_ and *I*(*t*) ≥ 0

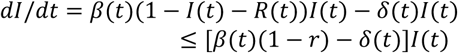

We consider discrete time steps in our modelling and measurements are taken on daily basis. Thus, replacing differential with difference equation.

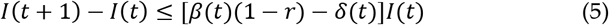

Solving for *I*(*t*)

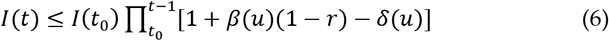

Since 0 ≤ *r* < 1

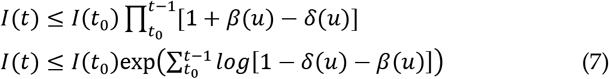

Expanding log as Taylor series and taking only the first term,

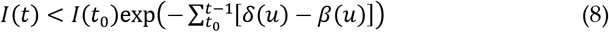

Considering a constant average di<u>ff</u>ere<u>n</u>ce between transmission rate and removal rate 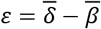 within the period *t*_0_ = 0 and *t* = *T* such that 0 ≤ *ε* ≤ 1

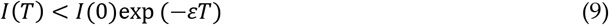

Considering *NI*(*T*) < 1 as disease free equilibrium state, the upper bound of *ε* can be derived as following such that the epidemic reaches baseline in time T.

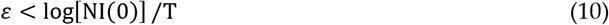

Maintaining *ε* > 0 asymptotically converges the total infection count to 0 at exponential rate thus makes the disease-free equilibrium stable.

Assuming a constant mortality rate, from (10) it can be deduced that increasing the recovery rate will directly reduce the time span of the disease outbreak. However, there is a hard limit for the removal rate, *δ*(*t*) ≤ 1. But *β*(*t*) can be greater than 1, specially during initial outbreak when total infection count is low. In such situation dampening the spread of infection will not be possible only with treatment facilities. Immediate restriction of mobility in area of outbreak and rapid isolation of infected individuals can reduce the transmission rate. Once it comes down below 1, enhanced treatment facilities can increase the recovery rate, thus reducing the span of the disease outbreak.

## 4 Spatiotemporal modelling of transmission rate

The transmission rate *β* can vary spatially as well as temporally based on multiple variables. Geographical location, weather conditions [13], human mobility [14], population statistics might be some of the impacting factors changing the dynamics of the spread. An exploratory analysis reveals probable dependency of multiple spatial and temporal features on the transmission rate. Spatially co-located regions might have similar dynamics of the spread with high autocorrelation of transmission rate in a localized region. However distant regions may have dissimilar transmission dynamics with low correlation. Thus, a large geographic area has been divided into small regions called as grids. Each grid has been divided even further into smaller regions called pixels. A population within a pixel is assumed to be constant and transmission dynamics is modeled by separate SIR models for each pixel. Each grid consists of co-located regions which might be impacting each other’s transmission rate. Feature is constructed for each grid as multichannel temporal sequence of matrices which in turn used for training a ConvLSTM [15] network to model the transmission rate. Data has been obtained for a region in United States from multiple sources [16, 29, 30, 31, 32, 33, 34]. The time span of the data is from 2020-03-21 to 2020-05-11.

### 4.1 Feature Construction

Covid19 daily data at USA county level are filtered by a spatial region of USA as shown in Fig. 1. The region is geospatially divided in M x N grids of equal sizes bounded by calculated latitudes and longitudes.

**Fig. 1.**
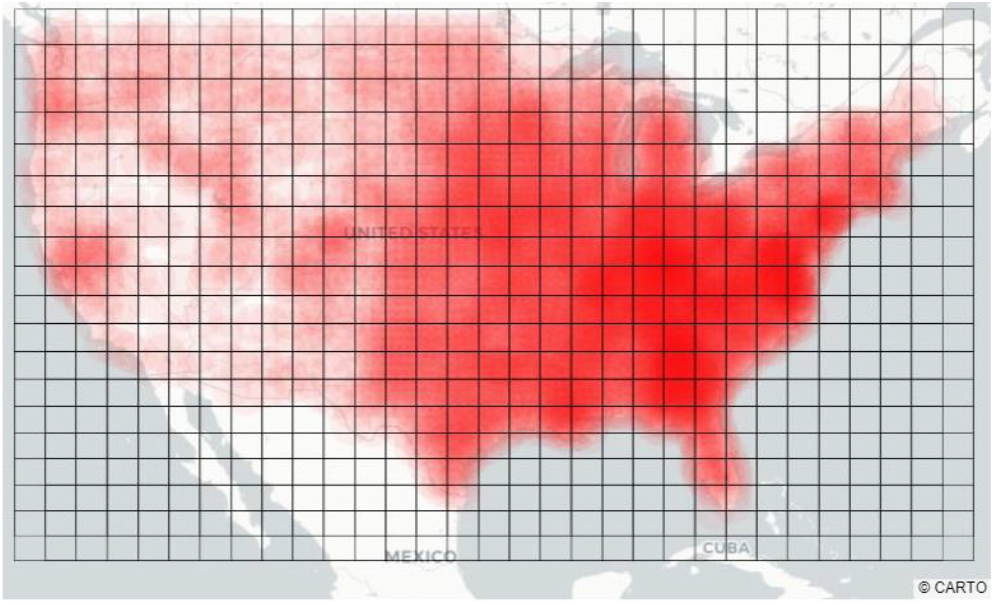
A region of USA divided in 18×30 grids. The red bubbles denote cumulative number of Covid-19 cases

Fig. 2a illustrates a grid bounded by latitudes and longitudes. The dotted line box is called as frame. The overlapping areas in all 4 directions in a frame allows flow of spatial influence from neighboring grids. A frame is in turn divided into L x L pixel. Each pixel represents a bounded area in geospatial region. Each pixel contains a value mapped to certain feature in the bounded geospatial region. Frame matrices are constructed for each feature and concatenated through a third axis called channels. For example, transmission rate and population density are two features and they represent two separate L x L matrices in a frame concatenated across a third axis. Some features like transmission rate, active infection fraction, weather etc. are distributed spatio temporally. Whereas other features like population density, female fraction, median age are assumed time invariant and have no temporal component. Thus, they are only distributed spatially and copied along temporal axis. Population density has been log transformed to reduce skewness and normalized. Other features are only normalized in 0-1 scale. Daily transmission rate and removal rates at pixel level have been calculated as following, where *i* ∈ {1.. *n*} denotes each pixel, 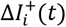 and Δ*R*_*i*_(*t*) are fraction of new cases in infected class and new individuals in removed class respectively at time *t* in pixel *i*.

**Fig. 2.**
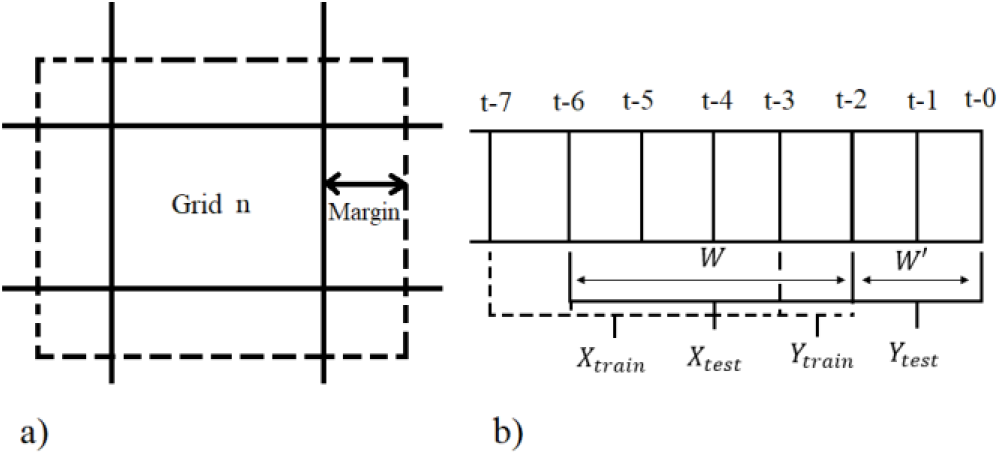
a) Illustration of overlapping frames obtained by spatially dividing a geographical region. The bold lines represent latitudes and longitudes which separates the grids. The box with dotted line represents the overlapping frame that is used for training the model. Each grid is divided LxL pixels. The margin refers to number of pixels of overlapping region. b) Illustration of sequence of a frame. t-0 is the most recent frame. X_train_, Y_train_ are the training samples and X_test_, Y_test_ are testing samples.

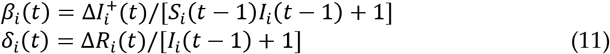

Each training sample of a frame is represented by a tensor of dimension T x L x L x C, where T is the total time span and C is number of channels or features. As shown in Fig. 2b each training sample is generated by sliding a time window size of W+1 by 1, leaving behind a test case sample of time window size of W^′^ in the most recent period. Number of training samples for a frame can be calculated as T − W^′^ − W − 1. Thus, total number of training samples *S*_*train*_ for all frames can be calculated as S_train_ = (T − W^′^ − W − 1) * M * N.

The forecasting problem is framed as supervised learning problem. Given a sequence of observed multichannel frames of spatial data as matrices *X*_1_, *X*_2_ … *X*_*t*_ the objective of the model is to predict the next single channel frame *Y*_*t*+1_. The training samples are divided into input sequences of length W and output frames. The model forecasts the transmission rate in each pixel in a frame for each timestep. Thus, the output frame consists of only 1 channel. The input training dataset (X_train_) can be represented as a tensor of size S_train_ x W x L x L x C and the output dataset (Y_train_) as S_train_ x W x L x L x 1. For training, the input sequences are selected from all frames having non-zero total infection count. Fig. 1b illustrates the sequence of a frame. The frames t-7 to t-3 represents an input training sequence (X_train_) of length W. The output frame (Y_train_) for this training sample is t-2. Other training samples are generated by sliding the window W+1 backwards in time by 1. The most recent images t-0 and t-1 represents the test output images (Y_test_) and immediate sequence of images t-6 to t-2 is the test input sample (X_test_). The test set X_test_ is represented by a tensor of size (M * N) x W x L x L x C and Y_test_ by (M * N) x W′ x L x L x 1.

### 4.2 Exploratory analysis of transmission rate

The primary purpose of the exploratory analysis is to understand the distribution of transmission rate and identify probable influence of different features on the transmission rate. Eight external features are analyzed against transmission rate to find probable influence. Among eight features, four are spatial features having no temporal component, namely population density, housing density, female fraction, median age. Fig. 3 illustrates scatter charts between average transmission rate and four spatial features for multiple pixels. The color gradient represents log transformed cumulative number of infection cases in each pixel. Only those pixels are filtered which experienced at least 30 days of running infection cases and having at least 10 cumulative infection cases at the beginning of the observation period. Fig 3a and 3b displays scatter charts and regression lines of average transmission rate with respect to population density and housing density in each pixel respectively. The two external features are log transformed and scaled to get upper bounded by 1. Log transformation reduces skewness and influence of outliers in data. As observed in the charts the transmission rate is positively correlated with both the features which is quite intuitive. Places with high population density is expected to experience rapid spread of the disease. Locations with high population density also experienced highest number of cumulative cases. Fig. 3c and 3b displays scatter charts and regression lines of transmission rate with respect to female fraction and median age of the population respectively. In Fig 3c, 16 pixels have been filtered out having female fraction less than 0.45 to remove the skewness in the data. There is a slight positive correlation between female fraction and transmission rate. However, this might not invoke a suggestive idea about the dependency of this external feature on transmission rate as majority of the pixels resides in the range of 0.50 – 0.53 female fraction with barely any trend in that range. Also, there is an indirect correlation as in general pixels with high female fraction has high population density. Median age has negative correlation with transmission rate. There is an indirect correlation in this case also as in general pixels with high median age has low population density. Another intuitive assumption can be, population with high median age are less mobile thus restricting the spread of the disease.

**Fig. 3.**
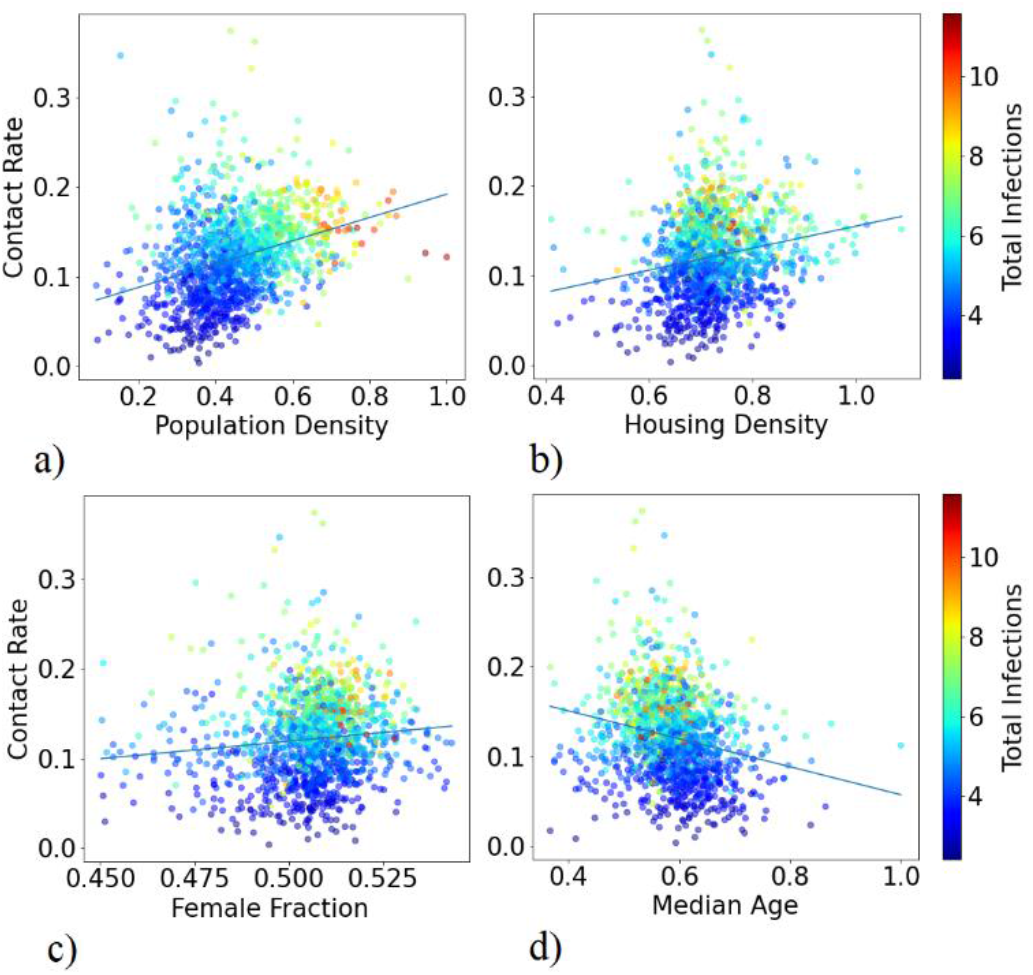
Average transmission rate (contact rate) in pixels and plotted against multiple features. The points are color coded based on normalized log transformed total patient count in each pixel. Plot of mean transmission rate against a) scaled log transformed population density, b) scaled log transformed housing density, c) fraction of female population, d) scaled median age

Apart from four spatial features four other external spatiotemporal features are analyzed to observe any influence on transmission rate. Fig. 4 illustrates time lagged cross correlation between transmission rate and other spatio-temporal features at pixel level. The external features are time lagged from 0 to 15 time steps and cross correlated with transmission rate for each lag. In the plot, pixels are arranged in increasing order of total infection cases. Fig. 4a and 4b shows the plot of cross correlation of transmission rate with respect to average daily temperature and 3-day running window temperature standard deviation respectively. Average temperature is slightly positively correlated in time lag range of 0-5. In the plot, offset 15 denotes time lag 0 and offset 0 as time lag 15. The correlation with temperature variation varies widely across pixels. However, on average there is a minor positive correlation in time lag range of 5-15. For both the features pixels having high total infections have negative correlation with transmission rate in the time lag range of 0-10. Fig. 4c and 4d shows the plot of cross correlation of transmission rate with respect to average daily relative humidity and daily removal rate respectively. There is an overall positive correlation with respect of relative humidity specially in pixels with highest infection cases. Removal rate is mostly negatively correlated with transmission rate except in few pixels having highest infection cases.

**Fig. 4.**
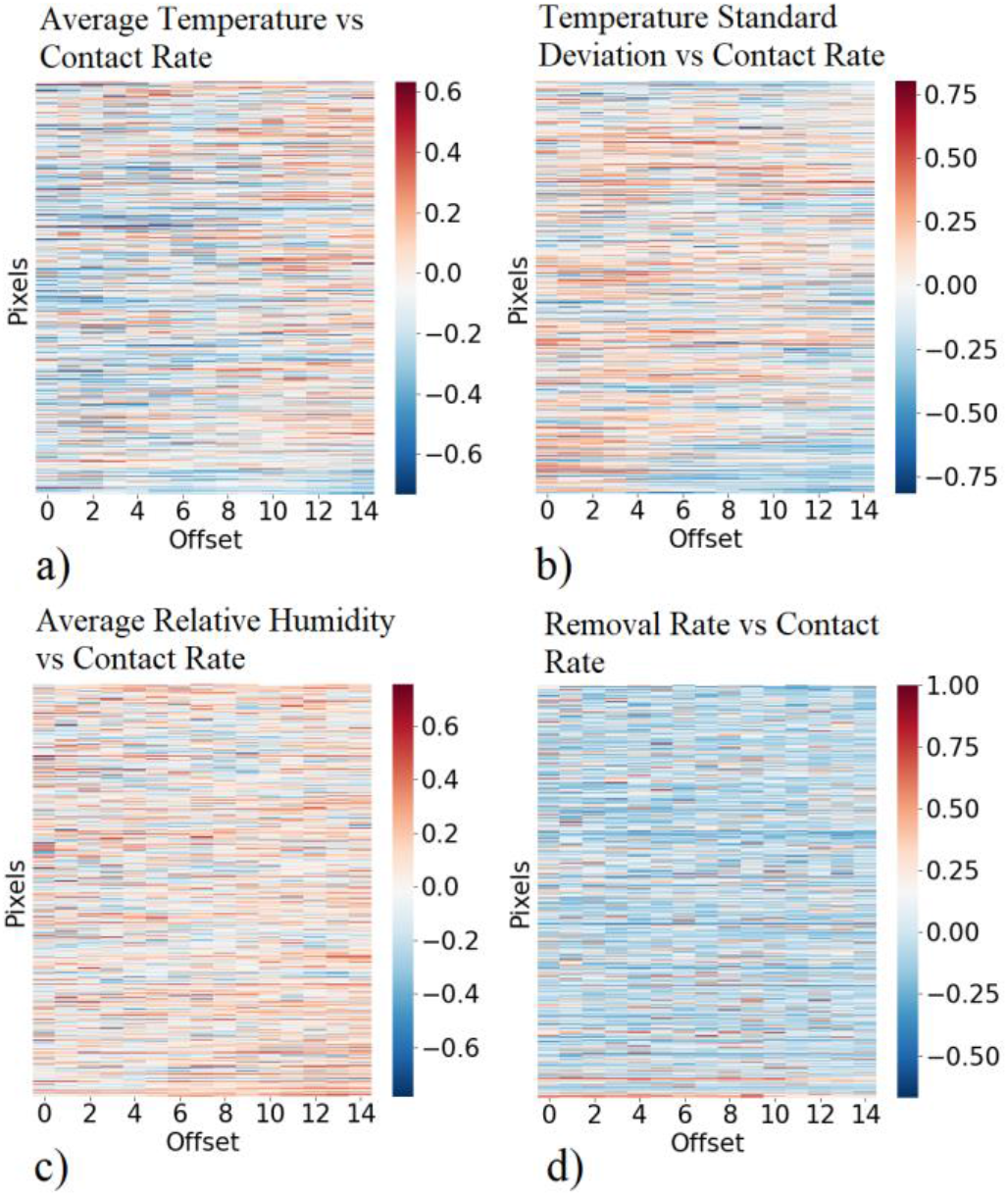
Time lagged Cross correlation between transmission rate (Contact rate) and different features across multiple pixels. Plot of cross correlation of transmission rate vs a) average temperature, b) temperature standard deviation in 3-day period, c) average relative humidity, d) removal rate

Correlation might not represent causality. Thus, we performed granger causality test [17] of transmission rate with respect to different features. Granger causality is a statistical hypothesis test for finding if one time series can help improving the forecasting accuracy of another time series. It might not measure true causality rather it measures predictive causality. Chi square test is chosen as the hypothesis testing method and minimum pvalues for each pixel are calculated. Augmented Dickey-Fuller test [18] is performed to test stationarity of all the timeseries. Table 1 displays the result of granger causality and Dickey-Fuller tests. The column ‘% of pvalue < 0.05’ represents percentage of pixels for which the granger causality test gave pvalue less than 0.05 for each feature. The column ‘% of ADF<10%’ represents percentage of pixels for which the Dickey-Fuller test gave test statistic less than 10% critical value and having pvalue less than 0.1 for each feature. From the observed results it seems for majority of the pixels the weather features and removal rate have predictive causal relation with transmission rate. Also, for majority of the pixels the feature timeseries are stationary or weakly stationary.

**TABLE 1.** Granger Causality test of Transmission Rate vs different Features

| Feature | % of Pvalue<0.05 | % of ADF<10% |
| --- | --- | --- |
| Transmission Rate | NA | 77.35 |
| Average Temperature | 74.19 | 67.26 |
| Temperature Standard Deviation | 76.82 | 71.93 |
| Average Relative Humidity | 73.66 | 86.38 |
| Removal Rate | 72.46 | 72.31 |

### 4.3 ConvLSTM model of transmission rate

Recurrent neural networks (RNN) are a class of artificial neural networks with nodes having feedback connections thereby allowing it to learn patterns in variable length temporal sequences. However, it becomes difficult to learn long term dependencies for traditional RNN due to vanishing gradient problem [19]. LSTMs [20] solve the problem of learning long term dependencies by introducing a specialized memory cell as recurrent unit. The cells can selectively remember and forget long term information in its cell state through some control gates. In convolutional LSTM [15] a convolution operator is added in state to state transition and input to state transition. All inputs, outputs and hidden states are represented by 3D tensors having 2 spatial dimensions and 1 temporal dimension. This allows the model to capture spatial correlation along with the temporal one. In our model we configured multichannel input such that distinct features can be passed through different channels. Multiple convolutional LSTM layers are stacked sequentially to form a network with high nonlinear representation. The final layer is a 2D convolutional layer having one filter which constructs a single channel output image as the next frame prediction. We assume the transmission rate saturates as number of infection cases increases. Thus, the modified transmission rate is calculated as 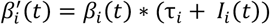 which serves as the response variable for the model and τ_*i*_ = 1/*N*_*i*_ and *N*_*i*_ is total population in pixel *i*.

The model is tested by feeding in input sequence of frames and next output frame is predicted which in turn is combined with other features along channel and appended with the input sequence. The new input sequence is fed to the model again to get the next predicted frame. This continues until forecasting completes for a desired time period. “Mean absolute percent error” (MAPE) and Kullback-Liebler (KL) divergence [17] are used to measure the accuracy of the model. The model predicts the transmission rate for a future time period for each pixel which in turn is used to calculate daily new infection cases 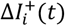 using equation 11. The removal rate is estimated as running average of previous 3-days and daily removed cases are calculated using equation 11. The active infection cases (*I*_*i*_ (*t*)) and susceptibles (*S*_*i*_(*t*)) are calculated using equation 1 and 2. Cumulative infection cases 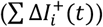 are calculated by summing up all new infection cases upto a certain day. MAPE of modified transmission rate is calculated at pixel level for the prediction period and averaged. The pixels with 0 susceptible population count are filtered out while calculating MAPE and KL divergence. Pixel MAPE is calculated as per equation 12, where G is set of all grids and G′ set of all pixels such that the frame for each corresponding grid have non zero cumulative infection count, *W*′ is prediction time period, *W*^′′^ = *T* − *W*′ is total time period in training set, 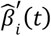 and 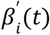 are predicted and actual modified transmission rate for *it*h pixel at time *t* respectively.

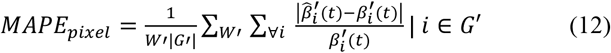

KL divergence at pixel level is calculated for modified transmission rate in the prediction period to measure the dissimilarity of distribution of predicted transmission rate with respect to actual. *σ* is softmax function applied after scaling a series in 0 to 1 scale and *P*(*X*) is probability distribution of *X*. Softmax is applied to convert total infection cases as probability distribution across pixels. Since KL divergence measures the dissimilarity between two distribution thus a lower value of it indicates better performance of the model.

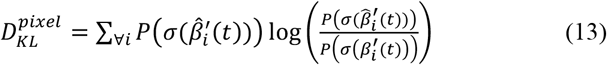

MAPE is also calculated at grid and country level with respect to cumulative predicted infection cases across the region during the prediction period.

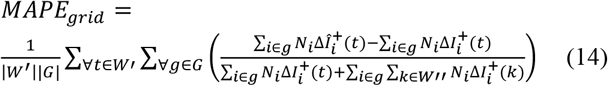

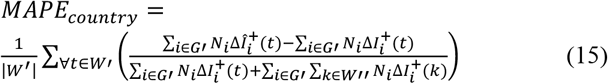

### 4.4 Model test results

The model is constructed by stacking 4 Convolutional LSTM layer sequentially and terminating the network with a Convolutional 2D layer. The final layer is followed by exponential linear unit as activation. The input and other hidden Convolutional LSTM layers are followed by sigmoid activation. Each Convolutional LSTM layer has 32 filters and kernel size 3×3. The input layer is configured to take tensors of size 16×16×8. Eight input features are constructed and fed into the model as separate channels. Namely transmission rate, population density, female fraction, median age, active infection fraction, average temperature, temperature standard deviation and average relative humidity. The model is trained for 20 epochs with batch size of 50 and mean squared error as loss function. Out of 11378 samples 10809 are used for training the model and 569 are for validation. The model is trained and tested twice. Once with all the eight features another with only five leaving out the weather features.

The dataset has a time span of 51 days out of which data from 42^nd^ to 51^st^ day is used for testing the model and rest for training and validation. Table 2 displays the training, validation and test results of the model. Statistics suggests there is a slight improvement of overall accuracy when weather features are included while training the model.

**TABLE 2.** Model training, validation and test results

| Metric | Value with weather features | Value without weather features |
| --- | --- | --- |
| Training mean absolute error | 0.0140 | 0.0140 |
| Validation mean absolute error | 0.0043 | 0.0063 |
| Pixel KL divergence | $8.306 \times 10^{-9}$ | $8.338 \times 10^{-9}$ |
| Pixel MAPE | 7.95% | 10.10% |
| Grid MAPE | 8.39% | 10.36% |
| Country MAPE | 0.19% | 0.25 % |
| Predicted total cases (1330525) | 1331175 | 1328605 |

Pixel MAPE and grid MAPE are below 10% in both the cases and country MAPE is below 1%. Predicted total infection cases at the end of prediction period is little overestimated than actual (1330525) when weather features are included in modelling and overestimated when weather features are not included. All future reference of trained model suggests the model has been trained with all eight features unless otherwise mentioned. Fig. 5 illustrates different plots of predicted vs actual infection cases in 10-day prediction period. Fig. 5a and 5b shows the plot of predicted vs actual new infection cases and cumulative infection cases per day in 10-day period. Fig. 5c and 5d shows the plot of predicted vs actual log transformed total new infection cases and cumulative infection cases per grid in 10-day prediction period. All the predicted curves closely approximate the actual values.

**Fig. 5.**
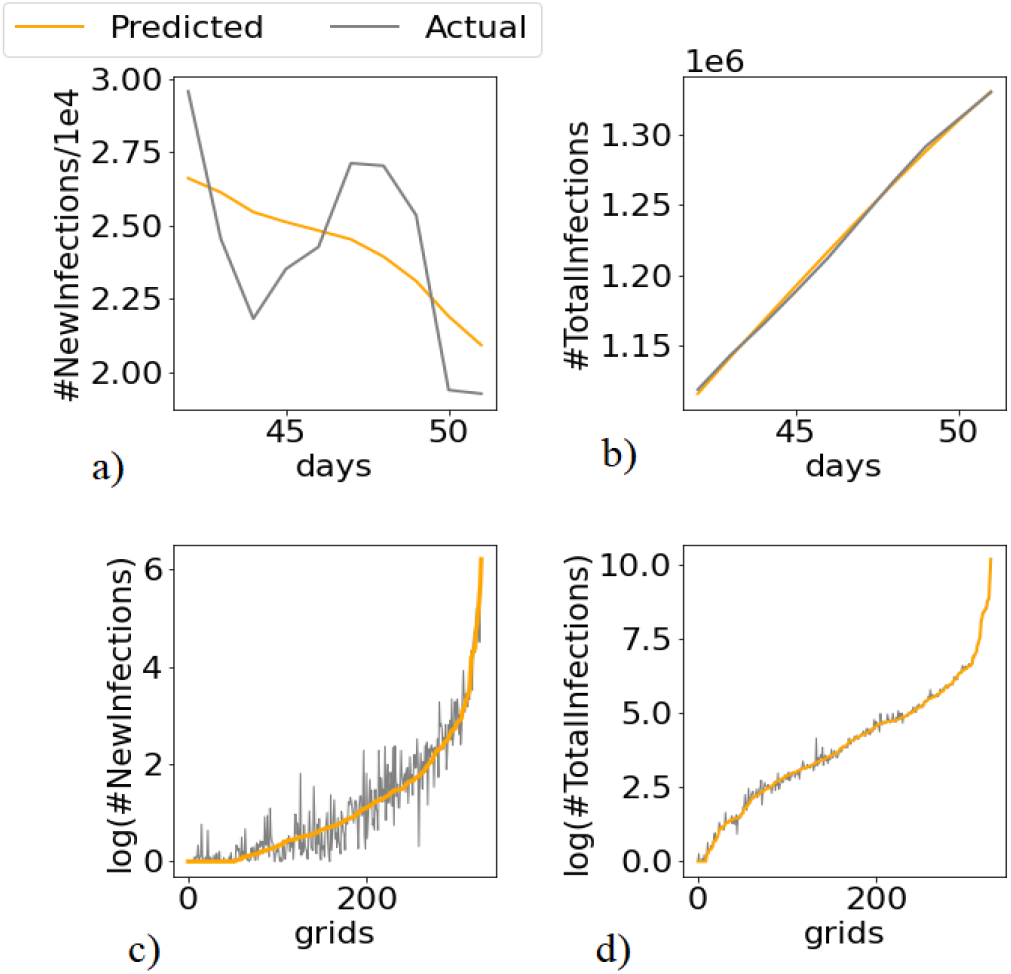
Predicted vs actual new and total infection count in a 10-day prediction period. Model is trained with all features. a) Days vs scaled new infection count across all pixels. b) Days vs scaled cumulative infection count across all pixels. c) Grids vs logarithm of sum of new infection count. d) Grids vs logarithm of sum of cumulative infection count.

## 5 Explaining influence on transmission rate

One of our goal of this study is to understand how different external features are influencing the transmission rate. We expect to find simple interpretable predictive causal relations between transmission rate and different features. One of the ways to find such relations is building an accurate predictive model followed by explaining the predictions in terms of input features. As described in previous sections deep neural networks can model the dynamics of epidemic quite accurately due to its high nonlinear representation. However high accuracy is tradeoff against model interpretability. Given the complexity of the Convolutional LSTM network used to model the transmission rate it is nearly impossible to find how each feature is influencing the transmission rate just by studying the weight matrices. Using a high bias predictive model like linear regression or shallow decision tree not only reduces the accuracy but also drops interpretability [21]. Simple models can serve as interpretable models but may fail to capture true relations among features globally. This problem can be solved by building simple local models and drawing local explanations of feature relations. However, there may not be enough data points available or data distribution may be highly skewed in a local region to confidently build a predictive model and draw interpretations on it. Thus, we use the trained Convolutional LSTM model as the global model and draw spatio-temporal local interpretations of it using locally perturbated synthetic data by satisfying a criterion called local fidelity [22]. Local fidelity suggests the explanations should be locally faithful with the model behavior. Local fidelity does not imply global fidelity however global fidelity implies local. To increase interpretability simple surrogate models can be trained with local data as it is expected that the response variable varies with the features almost linearly in a local region. In fact, there is a tradeoff between local fidelity and interpretability that needs to be made. Model agnostic methods perturbs the input features in a local region around a single or a group of datapoints and feeds the model to obtain predicted response variable. This synthetic data is in turn used to train simple surrogate models to obtain local interpretations of global model. There are several existing methods available in the literature to derive local interpretations of a model [22,23, 25]. Few works also proposed methods to derive global explanations from local interpretations of any black box models [21, 24].

### 5.1 Spatiotemporal locality of Transmission rate

Similar to as stated in [22] deriving explanations requires optimization of the following function, where G is set of interpretable surrogate models in a locality, *f* is the global model to be explained, *l*_*x*_ is the distribution function defining the locality of *x*, L is the loss function and *Ω*_*g*_ is the complexity of the model *g*.

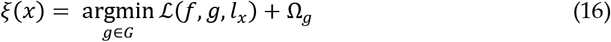

It is desirable to minimize both Ω_*g*_ and L. However, in general they are inversely proportional when the spread of *l* is large. A very small spread of *l*_*x*_ is also not desirable as it will oversimplify g to draw any meaningful explanations in the locality. Thus, a choice of *l*_*x*_ is important to derive meaningful interpretations.

The locality of *x* is defined by a threshold distance in all directions from *x* both spatially and temporally and it is defined by the following tuple, where *x*_*s*_ and *x*_*t*_ are spatial and temporal components of observation *x. d*_*s*_ and *d*_*T*_ are spatial and temporal threshold distances from *x* to the boundary of locality.

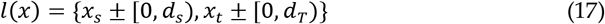

Fig. 6 illustrates spatiotemporal locality of observation *x*. Spatial locality is bounded by pixels up to *d*_*s*_ in all direction from *x*_*s*_ such that locality of *x*_*s*_ is bounded by a square box of pixels of size (2*d*_*s*_ + 1) X (2*d*_*s*_ + 1). No paddings are applied at the edges. Thus, perimeter defining locality of pixels at the edges of a frame are trimmed. As illustrated in Fig. 6 temporal locality is also defined similarly. Combining spatial and temporal locality the local region of observation *x* is defined by a sequence of group of pixels with equal time lead and lag from *x* unless *x* resides on temporal edge of an input tensor in which case temporal locality is trimmed on the direction of the edge.

**Fig. 6.**
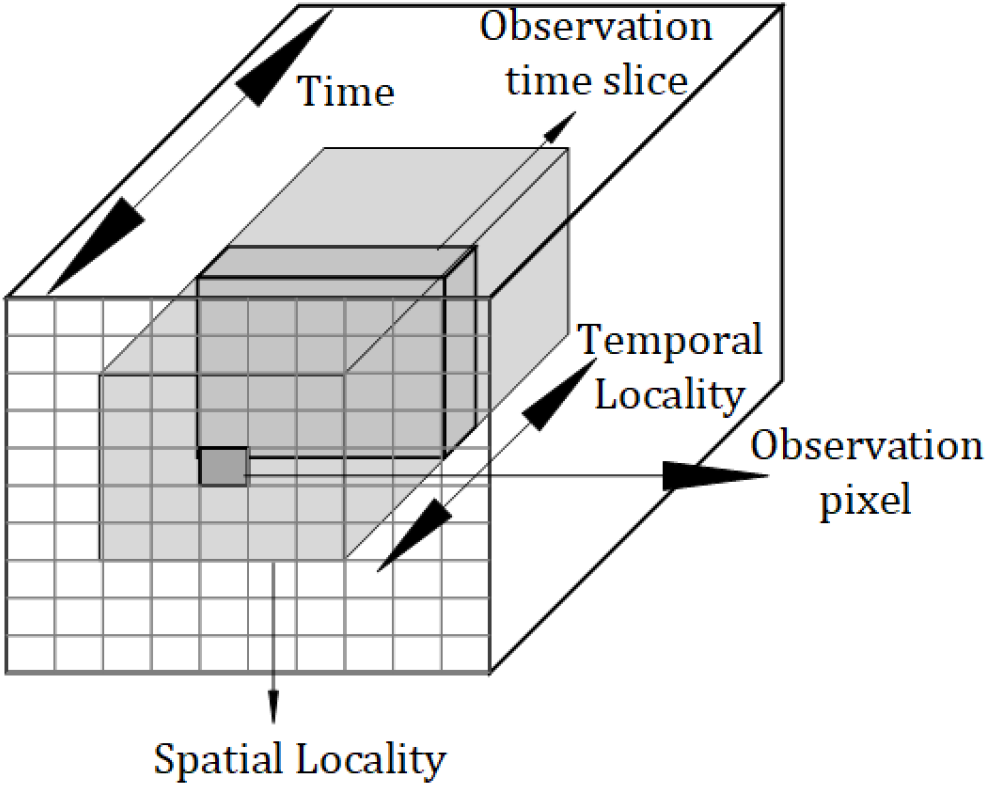
Illustration of spatio temporal locality

Perturbated data points are generated by randomly perturbing the pixel values of *x* following a uniform distribution. Perturbated distribution is calculated separately for each feature. The perturbated sample distribution is calculated as following, where *U*(*k, k*′) is uniform distribution with upper and lower bound as *k, k*′, *σ*(*l*(*x*)) is standard deviation of all observations in the locality of *x* and *rand* randomly selects one sample from two.

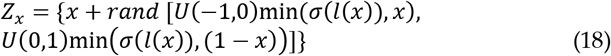

The spatial features are only perturbated spatially and same values are copied temporally along the corresponding channel. The channels having temporal component are perturbated for different time slice within an input tensor. Each perturbated pixel in a time slice represents a separate feature. Input tensors are constructed using the perturbated values and passed through the blackbox model *f* to generate a predicted output value. The set of all input perturbated data points of *x* and the corresponding predicted output values serves as the training dataset for the surrogate model *g*. Each input channels and the predicted values are normalized to 0 mean and 1 standard deviation prior to training the surrogate model. Normalization is done to convert the features into same scale so that coefficients of a linear regression surrogate model gives the relative influence of the features on the response variable. Thus, the loss function is defined as following, where the function *S* constructs the input tensor in the original representation from perturbated samples.

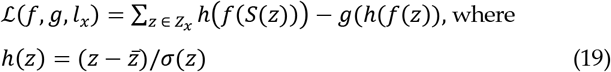

Though *Z*_*x*_ can be created by perturbing all features of a pixel in each channel within an input tensor, however in our analysis only a subset of all features is perturbated to produce *Z*_*x*_ to find effect of those features on transmission rate. Other features are kept constant as per the original observation. Intuitively this will explain the effect of the chosen features on the transmission rate in a single pixel area given all other parameters remain constant including feature values of spatio temporal neighboring pixels.

### 5.2 Interpreting prediction of Transmission rate

In our analysis *Z*_*x*_ is created by perturbing the following features only. Population density, female fraction, median age, weather at 4^th^, 5th and 6^th^ time lag. Weather includes average daily temperature, 3-day temperature standard deviation and average daily relative humidity. Apart from the weather features the other three features have no temporal component. So, for them the perturbated values are copied temporally in the input tensor during reconstruction. The weather features from 4^th^ to 6^th^ time lag is chosen by assuming the average incubation period of Sars-Cov-2 between 4 to 6 days. The spatial (*d*_*s*_) and temporal (*d*_*T*_) distance for defining locality is taken as 1. The perturbated samples for each feature are generated by equation 18. Local interpretations are carried out for each pixel which experienced at least 10 cumulative infection cases on 21^st^ March 2020. The objective is to deduce the influence of aforementioned features on the transmission rate in each pixel given all other parameters remains constant. 250 perturbated input samples are generated for each pixel. The samples are reconstructed in tensor format and fed to the model to obtain the predicted transmission rate and together they form the input output samples. For each pixel a linear regression surrogate model is trained with the training samples. The coefficients of each feature denote the influence on the transmission rate.

Fig. 7 illustrated the feature influence chart for different pixels in grid 387. We choose grid 387 as it experienced highest number of cumulative infection cases with nearly 10% of total infection cases in USA as of 1^st^ May 2020. Only those coefficients are plotted which have pvalue < 0.05. The features whose absolute value of median and standard deviation across all days are less than 0.05, are considered unimportant and filtered out from the plot. The counties covered by each pixel in grid 387 which have nonzero population is stated in Table 3. The Influence values are smoothed using 3^rd^ degree polynomial. New York & Bronx have somewhat positive influence of population density (pop den) and female fraction (f perc) on transmission rate. Median age (med age) has positive effect in the mid period and negative on early and later days. 6^th^ day time lag temperature (T6 temp) have slight negative effect on later days. On average Putnam also have positive influence of population density, median age and female fraction. However, population density and female fraction shows negative influence on later days. 4^th^ time lag and 5^th^ time lag relative humidity (T4 rh & T5 rh) have slight negative impact on average. At grid level population density and female fraction positively impacts transmission rate on daily basis. Median age has minor positive impact on earlier days and negative impact on later days. Fig 7d. shows median of influence across all days for different pixels in grid 387. Population density and female fraction have positive impact across all pixels. Median age closely resembles a sinusoidal curve which implies that its influence varies widely across pixels.

**TABLE 3.** County names for each pixel in grid 387

| Pixel | County | Pixel | County | Pixel | County |
| --- | --- | --- | --- | --- | --- |
| 85 | King & Queens | 133 | Rockland | 166 | Putnam |
| 101 | New York & Bronx | 134 | Westchester | 171 | Middlesex |
| 102 | Nassau | 151 | Fairfield | 182 | Dutchess |
| 122 | Suffolk | 153 | New Haven | 184 | Litchfield |

**Fig. 7.**
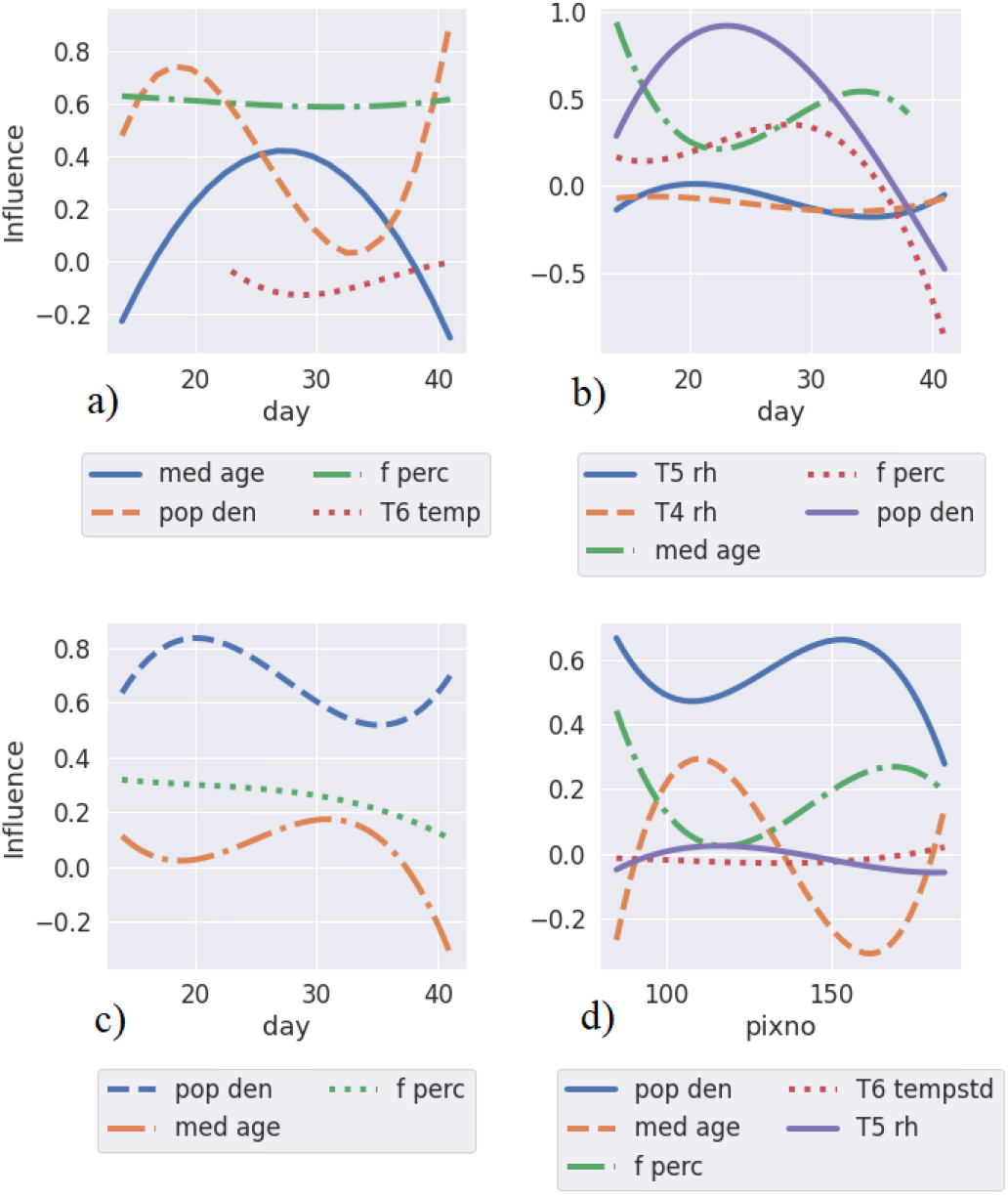
Feature influence chart by day, for pixels in grid 387. a) Influence chart of pixel covering the area of New York and Bronx, b) influence chart of pixel covering the area of Putnam, c) median influence chart by day for grid 387, d) median influence chart per pixel in grid 387

Fig. 8 illustrates the global effects of the features on transmission rate. To generate global interpretations local surrogate models are built for each pixel with 100 perturbated samples. For each feature the distribution of influence values for all pixels with nonzero population is plotted against time. Considering the median of the distribution, population density, female fraction has positive impact across all days whereas median age has negative impact. Temperature has minor positive impact, temperature standard deviation has minor negative impact and relative humidity barely have any noticeable impact on transmission rate. From this study it is clear local influence of features at pixel and grid level may widely deviate from global average. This is important as spread of infection is highly skewed regionally such that few hotspots contribute majority of the infection cases. Thus, studying the local influence of features can shed light on the local dynamics of spread and at the same time global influence charts provides a general idea of the influence on spread.

**Fig. 8.**
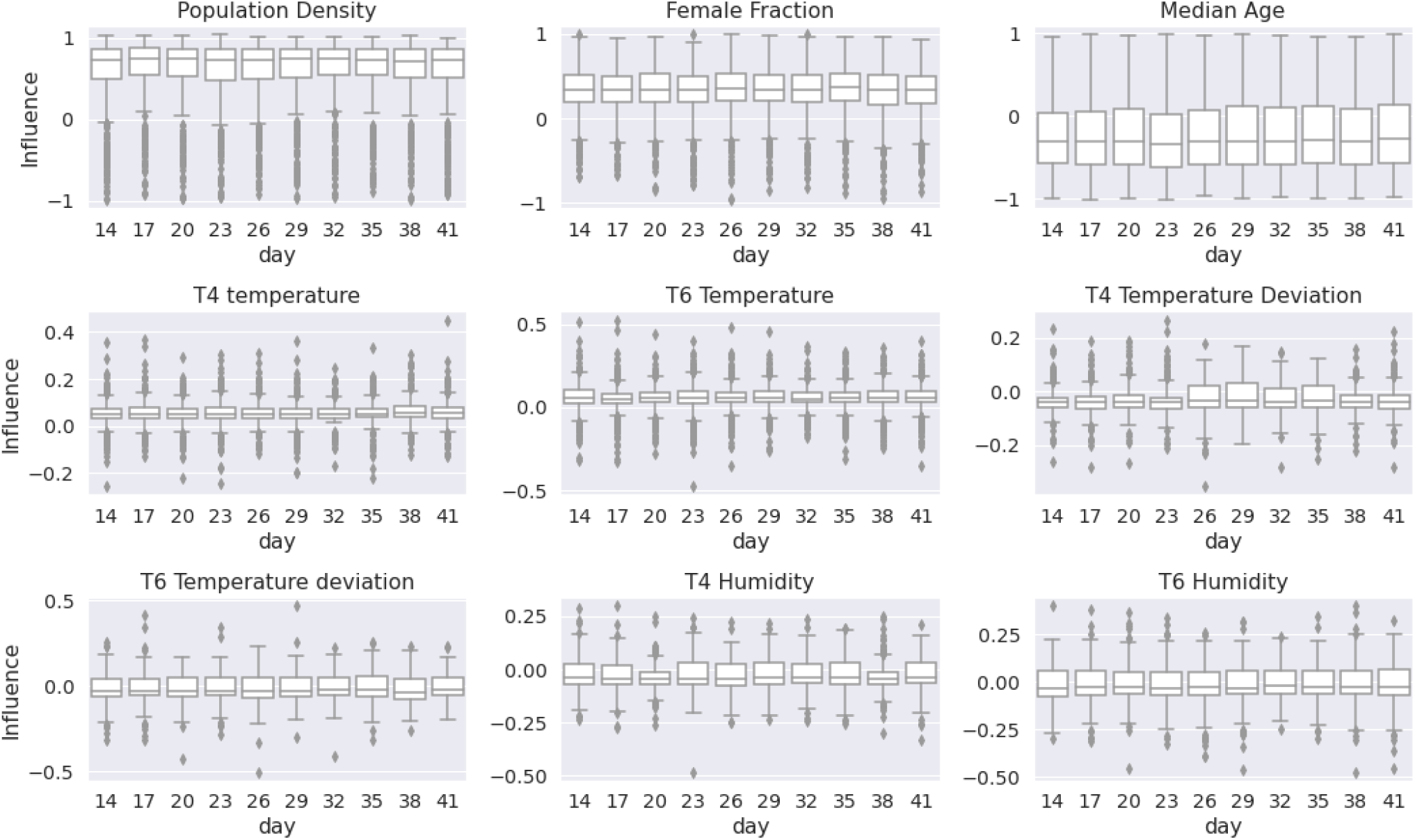
Global Influence of different features on transmission rate

## 5 Long term forecasting of disease progression

Classical SIR model assumes a constant transmission rate and it typically predicts a smooth bell curve of active infection cases with respect to time with a single peak. However, transmission rate may vary with respect to multiple external factors including intervention methods like lockdown. A variable transmission rate may result in periodic subsidence and resurgence of the spread of infection and in turn producing multiple peaks of active infection cases along time. Along with this the recovery rate may also change due to multiple intervention methods like enhancing hospital facilities, improving treatment procedure etc. As shown in equation 10, recovery rate is very important in achieving disease free stable equilibrium state. In general, the average removal rate (recovery rate + death rate) over a period should exceed average transmission rate in order to reach the disease-free equilibrium. Considering the death rate to be constant and quite small compared to recovery rate of Covid-19, the time required to reach the equilibrium state is inversely proportional to the difference between recovery rate and transmission rate. In our experiments we used the trained model to do long term forecasting of the epidemic with current normal parameters and compared with an “what if” analysis by modifying the removal rate.

A 300 days forecasting is carried out for the grid 387. Since weather features barely impacts transmission rate in grid 387 thus the model trained without weather features is used for forecasting. “What if” analysis is done by setting high removal rate to expedite disease-free equilibrium and compared with current normal forecasting by setting removal rate as running average of past 5 days. In “what if” analysis removal rate is set as per equation 10 by setting T = 200 with upper hard limit 0.2. As removal rate changes daily active infection cases which in turn impacts future transmission rate and due to upper hard limit of removal rate the value of *ε* in some pixels is less than upper bound calculated by equation 10. From fig. 9a and 9b it is evident that number of active infection cases reduced much faster in the “what if” analysis and most of the pixels hit near baseline state at least once within 200-day period. However rapid periodic resurgence of the disease is seen in this case. As recovery rate has upper hard limit thus in some cases resurgence with high transmission rate resulted in destabilizing disease-free equilibrium. The growth is again quickly dampened due to high recovery rate in future periodic resurgences. This can be empirically explained by the fact that population gets cautious and maintains social distancing with low intermixing when infection cases are high and vice versa. Fig. 9c and 9d suggests there is rapid periodic resurgence of new infection cases in “what if” analysis compared to current normal and multiple short low new infection periods are seen. The resurgences in some cases (pixel 85, 101) are stronger compared to current normal. Thus, it is evident, by only increasing recovery rate abruptly, infection spread may not be controlled fully unless other intervention methods are adopted to prevent spike of transmission rate during resurgence periods. Fig. 10a and 10b shows the plot of daily active infections when only pixel 101 and 102 are subjected to modified recovery rate respectively and other pixels are set with current normal recovery rate. In both the cases there is a quick dampening of active cases in 101 and 102 pixels and resurgence spike is shorter and weaker compared to Fig. 9b. It is evident there is spatial influence of neighboring active cases and transmission rate. One explanation can be, isolated intervention measures to dampen the spread does not breaks the cautiousness and preventive measures among the population. This makes determining an ideal recovery rate for a region a complex optimization problem.

**Fig. 9.**
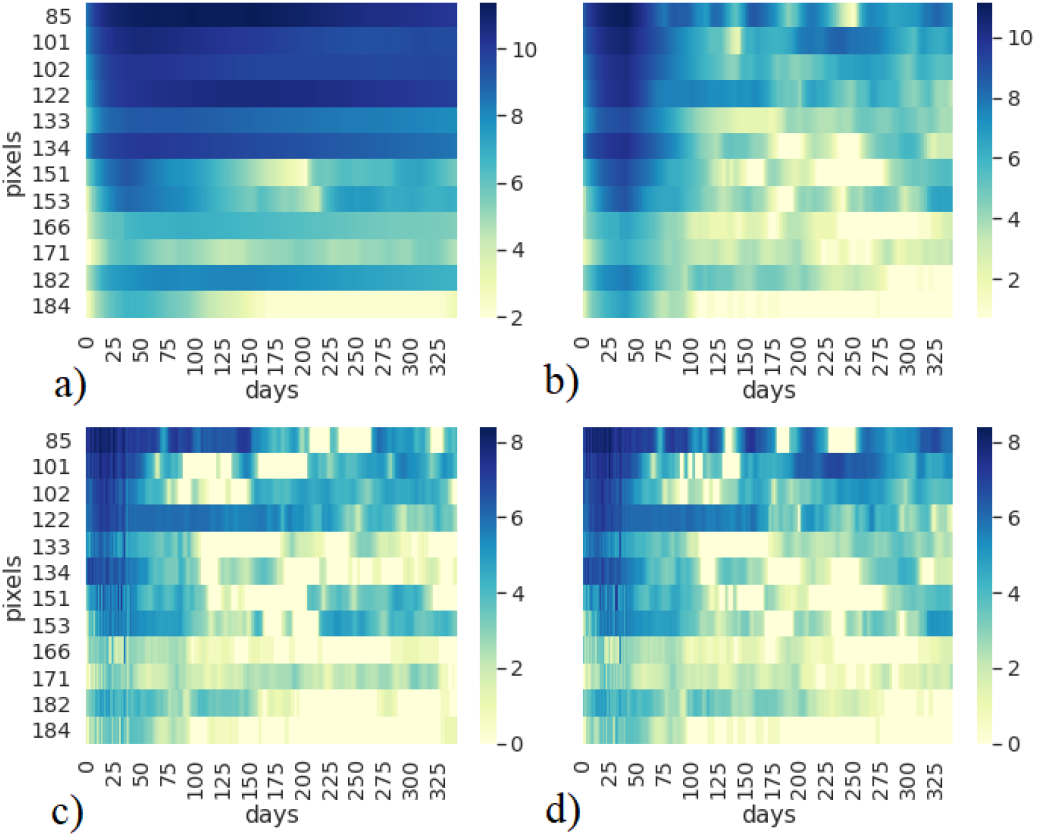
Plot of daily log transformed active infection cases and new infection cases for each pixel in grid 387. a) Daily active infections with normal running average removal rate b) daily active infections with modified removal rate to expedite disease-free equilibrium, c) daily new infections per pixel with normal running average removal rate d) daily active infections with modified removal rate

**Fig. 10.**
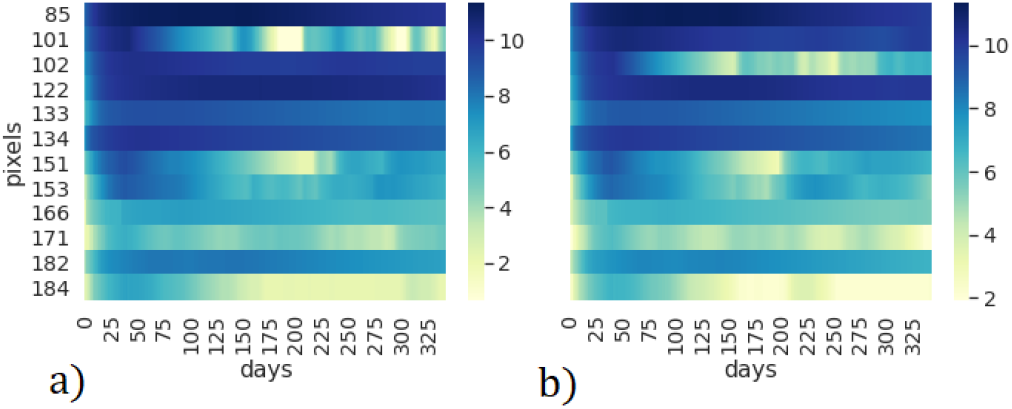
“What if” analysis with increased recovery rate for specific pixels in grid 387. Daily active infections with normal running average removal rate for all pixels except a) pixel 101 and b) pixel 102, where recovery rate is set to a high value in the forecast period.

Fig. 11 shows active infection cases at grid level quickly reaches baseline in “what if” scenario compared to current normal, but it is not eradicated fully. There are also small periodic spikes in future. The current normal scenario suggests unless strict intervention actions are not taken to reduce transmission rate or recovery rate it is going to take long time to reach the baseline. The trace of new infection cases suggests the trend is quite similar in both the scenarios with more frequent and stronger spikes in “what if” scenario. In current normal scenario the model estimates 487254 new infection cases and 711040 removed cases in 300-day period. In “what if” scenario it estimates 549158 new infection cases and 909437 removed cases. However, Fig. 11c suggests most of the removal happens in initial 50 days of forecast period due to abrupt increase of removal rate in forecast period. In real world such abrupt increase of removal rate may not be possible. However, on an average if the difference between removal rate and transmission rate can be maintained as per equation 10 it is possible to dampen the spread of infection within desired time period. Though in our analysis we took removal in strict sense however it may not refer to complete recovery. Identification and complete isolation of a patient such that there is negligible chance of further spread of the infection from the patient may also be referred to removal. Thus, maintaining high recovery rate, rapid and strict isolation of infected patient and intervention methods to reduce transmission rate are the keys to rapid convergence to diseasefree equilibrium.

**Fig. 11.**
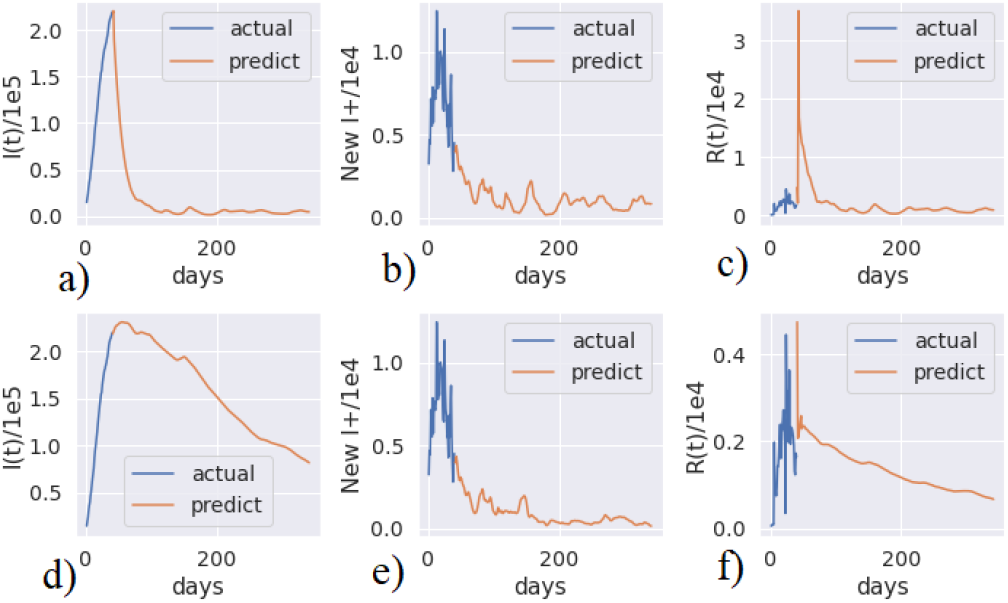
Comparison of forecasted infection spread between current normal scenario and “what if” scenario with increased recovery rate for grid 387. Trace of a) active infection cases, b) new infection cases and c) removed infection cases in “what if” scenario. Trace of a) active infection cases, b) new infection cases and c) removed infection cases in current normal scenario.

## 6 Conclusion

A thorough study on the transmission rate of Covid19 in USA revealed several insights. Key influencers are identified. However, there might be other influencers like human mobility, demographics, government interventions etc. On availability of those feature data, proposed methods may be applied to find influences. These methods can also be applied to other countries. Though a threshold condition is derived for disease free equilibrium, yet it is not straight-forward to determine ideal recovery rate to rapidly dampen the infection spread due to complex dependency of transmission rate. A general solution method may be investigated to solve this optimization problem and come up with ideal regional recovery rate.

## Data Availability

Data is available publicly and cited in the article

## References

[1] Kermack, W. O., & McKendrick, A. G. (1927). A contribution to the mathematical theory of epidemics. Proceedings of the royal society of london. Series A, Containing papers of a mathematical and physical character, 115(772), 700–721.

[2] Hethcote, H. W. (1976). Qualitative analyses of communicable disease models. Mathematical biosciences, 28(3-4), 335–356.

[3] Liu, W. M., Levin, S. A., & Iwasa, Y. (1986). Influence of nonlinear incidence rates upon the behavior of SIRS epidemiological models. Journal of mathematical biology, 23(2), 187–204.

[4] L. S. Chen and J. Chen, Nonlinear Biological Dynamics System, Scientific Press, China, 1993.

[5] Wei, C., & Chen, L. (2008). A delayed epidemic model with pulse vaccination. Discrete Dynamics in Nature and Society, 2008.

[6] R. M. Anderson and R. M. May, “Regulation and stability of host-parasite population interactions: I. Regulatory processes,” The SWARNA ET AL.: ON NONLINEAR INCIDENCE RATE OF COVID-19 11 Journal of Animal Ecology, vol. 47, no. 1, pp. 219–267, 1978

[7] Zhang, J. Z., Jin, Z., Liu, Q. X., & Zhang, Z. Y. (2008). Analysis of a delayed SIR model with nonlinear incidence rate.Discrete Dynamics in Nature and Society, 2008.)

[8] Xue, Y. A. K. U. I., & Duan, X. (2011). Dynamic analysis of an SIR epidemic model with nonlinear incidence rate and double delays. Int J Inf Syst Sci, 7(1), 92–102.

[9] P. van den Driessche, J. Watmough, A simple sis epidemic model with a backward bifurcation, Math. Biol. 40 (2000) 525–540.

[10] Liu, X., & Stechlinski, P. (2012). Infectious disease models with time-varying parameters and general nonlinear incidence rate. Applied Mathematical Modelling, 36(5), 1974–1994.

[11] Bacaër, N., & Gomes, M. G. M. (2009). On the final size of epidemics with seasonality. Bulletin of mathematical biology, 71(8), 1954.

[12] Hong, H. G., & Li, Y. (2020). Estimation of time-varying transmission and removal rates underlying epidemiological processes: a new statistical tool for the COVID-19 pandemic. arXiv preprint 2004.05730.

[13] Sajadi, M. M., Habibzadeh, P., Vintzileos, A., Shokouhi, S., Miralles-Wilhelm, F., & Amoroso, A. (2020). “Temperature and latitude analysis to predict potential spread and seasonality for covid-19”. Available at SSRN 3550308

[14] Zhan, C., Tse, C., Fu, Y., Lai, Z., & Zhang, H. (2020). “Modelling and Prediction of the 2019 Coronavirus Disease Spreading in China Incorporating Human Migration Data”. Available at SSRN 3546051

[15] Xingjian, S. H. I., Chen, Z., Wang, H., Yeung, D. Y., Wong, W. K., & Woo, W. C. (2015). “Convolutional LSTM network: A machine learning approach for precipitation nowcasting”. In Advances in neural information processing systems (pp. 802–810

[16] China Data Lab, 2020, “US COVID-19 Daily Cases with Base- map”,https://doi.org/10.7910/DVN/HIDLTK,HarvardDataverse,V18,UNF:6:s0u1J15PWisF3mouUiT6Kw==[fileUNF]

[17] Granger, C. W. (1969). Investigating causal relations by econometric models and cross-spectral methods. Econometrica: journal of the Econometric Society, 424–438.

[18] Dickey, D. A., & Fuller, W. A. (1979). Distribution of the estimators for autoregressive time series with a unit root. Journal of the American statistical association, 74(366a), 427–431.

[19] Hochreiter, S. (1998). “The vanishing gradient problem during learning recurrent neural nets and problem solutions”. International Journal of Uncertainty, Fuzziness and Knowledge-Based Systems, 6(02), 107–116.

[20] Hochreiter, S., & Schmidhuber, J. (1997). “LSTM can solve hard long time lag problems”. In Advances in neural information processing systems (pp. 473–479).

[21] Lundberg, S. M., Erion, G., Chen, H., DeGrave, A., Prutkin, J. M., Nair, B., … & Lee, S. I. (2020). From local explanations to global understanding with explainable AI for trees. Nature machine intelligence, 2(1), 2522–5839.

[22] Ribeiro, M. T., Singh, S., & Guestrin, C. (2016, August). “rWhy should i trust you?” Explaining the predictions of any classifier. In Proceedings of the 22nd ACM SIGKDD international conference on knowledge discovery and data mining (pp. 1135–1144).

[23] David Baehrens, Timon Schroeter, Stefan Harmeling, Motoaki Kawanabe, Katja Hansen, and Klaus-Robert MÃžller. How to explain individual classification decisions. Journal of Machine Learning Research, 11(Jun):1803–1831, 2010.

[24] Ribeiro, M. T., Singh, S., and Guestrin, C. Anchors: Highprecision model-agnostic explanations. In AAAI, 2018a.

[25] Plumb, G., Molitor, D., & Talwalkar, A. S. (2018). Model agnostic supervised local explanations. In Advances in Neural Information Processing Systems (pp. 2515–2524)

[26] Hu, Z., Ge, Q., Jin, L., & Xiong, M. (2020). “Artificial intelligence forecasting of covid-19 in china”. arXiv preprint 2002.07112.

[27] Fanelli, D., & Piazza, F. (2020). “Analysis and forecast of COVID- 19 spreading in China, Italy and France”. Chaos, Solitons & Fractals, 134, 109761.

[28] Xi, G., Yin, L., Li, Y., & Mei, S. (2018, November). “A deep residual network integrating spatial-temporal properties to predict influenza trends at an intra-urban scale”. In Proceedings of the 2nd ACM SIGSPATIAL International Workshop on AI for Geographic Knowledge Discovery (pp. 19–28). https://covidtracking.com/api/v1/states/daily.csv

[29] China Data Lab, 2020, “US COVID-19 Daily Cases with Base-map”, https://doi.org/10.7910/DVN/HIDLTK,HarvardDataverse,V24,UNF:6:4iyX7x/Oqi+dloA11aGvdQ==[fileUNF]

[30] U.S. Department of Commerce, Office of Textiles and Apparel. (2002). U.S. Total Exports in U.S. Dollars. Retrieved from http://otexa.ita.doc.gov/tqexp/htsdata.

[31] Menne, M.J., I. Durre, R.S. Vose, B.E. Gleason, and T.G. Houston, 2012: An overview of the Global Historical Climatology Network-Daily Database. Journal of Atmospheric and Oceanic Technology, 29, 897–910, doi:10.1175/JTECH-D-11-00103.1.

[32] Diamond, H. J., T. R. Karl, M. A. Palecki, C. B. Baker, J. E. Bell, R. D. Leeper, D. R. Easterling, J. H. Lawrimore, T. P. Meyers, M. R. Helfert, G. Goodge, and P. W. Thorne, 2013: U.S. Climate Reference Network after one decade of operations: status and assessment. Bull. Amer. Meteor. Soc., 94, 489–498.

[33] https://www.kaggle.com/headsortails/covid19-us-county-jhu-data-demographics?select=us_county.csv

[34] Paul S.K, Jana, S., & Bhaumik, P. (2020). A multivariate spatio- temporal spread model of COVID-19 using ensemble of ConvLSTM networks. medRxiv.

